# The association between diabetes and mortality among patients hospitalized with COVID-19: Cohort Study of Hospitalized Adults in Ontario, Canada and Copenhagen, Denmark

**DOI:** 10.1101/2022.07.04.22276207

**Authors:** Orly Bogler, Afsaneh Raissi, Michael Colacci, Andrea Beaman, Tor Biering-Sørensen, Alex Cressman, Allan Detsky, Alexi Gosset, Mats Højbjerg Lassen, Chris Kandel, Yaariv Khaykin, David Barbosa, Lauren Lapointe Shaw, Derek R. MacFadden, Alexander Pearson, Bruce Perkins, Kenneth J. Rothman, Kristoffer Grundtvig Skaarup, Rachael Weagle, Chris Yarnell, Michelle Sholzberg, Bena Hodzic-Santor, Erik Lovblom, Jonathan Zipursky, Kieran L. Quinn, Mike Fralick

## Abstract

**Importance:** Diabetes has been reported to be associated with an increased risk of death among patients with COVID-19. However, available studies lack detail on COVID illness severity and measurement of relevant comorbidities.

**Design, Setting, and Participants:** We conducted a multicenter, retrospective cohort study of patients over the age of 18 years who were hospitalized with COVID-19 between January 1, 2020 and November 30, 2020 in Ontario, Canada and Copenhagen, Denmark. Chart abstraction emphasizing co-morbidities and disease severity was performed by trained research personnel. The association between diabetes and death was measured using Poissson regression.

**Main Outcomes and Measures:** within hospital 30-day risk of death.

**Results:** Our study included 1018 hospitalized patients with COVID-19 in Ontario and 305 in Denmark, of whom 405 and 75 patients respectively had pre-existing diabetes. In both Ontario and Denmark, patients with diabetes were more likely to be older, have chronic kidney disease, cardiovascular disease, higher troponin levels, and to receive antibiotics compared with adults who did not have diabetes. In Ontario, the crude mortality rate ratio among patients with diabetes was 1.60 [1.24 – 2.07 95% CI] and in the adjusted regression model was 1.19 [0.86 – 1.66 95% CI]. In Denmark, the crude mortality rate ratio among patients with diabetes was 1.27 (0.68 – 2.36 95% CI) and in the adjusted model was 0.87 (0.49 – 1.54 95% CI)]. Meta-analyzing the two rate ratios from each region resulted in a crude mortality rate ratio of 1.55 (95% CI 1.22,1.96) and an adjusted mortality rate ratio of 1.11 (95% CI 0.84, 1.47).

**Conclusions:** Presence of diabetes was not strongly associated with in-hospital COVID mortality independent of illness severity and other comorbidities.

## INTRODUCTION

Since January 2020, over 450 million people worldwide have been infected with the Severe Acute Respiratory Syndrome Coronavirus 2 (SARS-CoV2), and more than 5 million people have died. Reported risk factors for contracting Coronavirus Disease 2019 (COVID-19) include male sex, older age, living in congregate settings such as a long-term care facility or shelter, or having social, economic, or personal barriers that limit healthcare access (1, 2). Reported risk factors associated with experiencing a more severe course of COVID-19 include older age, male sex, diabetes mellitus, hypertension, obesity, cardiac disease, and chronic kidney disease (3-5).

Much attention has focused on diabetes and COVID-19 given its reported association with a worse prognosis, including an increased risk of intensive care unit (ICU) admission (3, 6-9), mechanical ventilation (3, 10), and mortality (4, 11-14). Within these studies, there is considerable variability in the measured strength of the association between diabetes and adverse clinical outcomes. Uncertainty remains about whether diabetes itself increases the risk of adverse outcomes given its inherent pro-inflammatory state, or whether the association is a result of related comorbid conditions (given that diabetes is an independent predictor of microvascular and macrovascular disease) or differences in care processes (15-17). From a patient perspective, it is not known if someone with diabetes, but without complications or comorbidities, carries a comparable risk of severe clinical outcomes from COVID-19 as a person with diabetes and late-stage complications such as advanced kidney or cardiovascular disease.

Existing studies describing the relationship between diabetes and COVID-19 severity have typically included single center or regional data and patients with varying illness severity. The main objective of our study was to understand the association between diabetes and death among patients hospitalized with COVID-19.

## METHODS

### Study Setting and Data Source

We conducted a retrospective (Ontario) and prospective (Denmark) cohort study at 10 hospitals in Ontario, Canada and 8 in Copenhagen, Denmark. The sites represented a convenience sample of hospitals where the study team members worked. The electronic medical record was the primary source of data collection and manual data collection was performed by trained research personnel. Our study was approved by the institutional research ethics board at each participating institution. For data privacy reasons we were unable to combine the data from Ontario and Denmark into one single dataset and thus we report results separately.

### Study population

The cohort included adults 18 years or older hospitalized with COVID-19 between January 1, 2020, and November 30, 2020. Consecutive patients hospitalized with COVID-19 were identified using Infection Prevention and Control departments at each hospital and through access to a central repository where SARS-CoV-2 laboratory results were reported. Patients were classified as having diabetes mellitus by one of the following at hospital presentation: hemoglobin A1C ≥ 6.5%, current use of at least one oral or injectable diabetes medication, or chart review with a physician’s note indicating the patient had diabetes. We were not able to distinguish between type 1 and type 2 diabetes or classify diabetes by severity.

### Data Collection

Information abstracted included patient demographics (age, sex, English proficiency, and place of residence before the hospitalization: home, homeless, long-term care, transfer from another hospital), pre-existing medical conditions previously shown to be associated with mortality from COVID-19 (cardiovascular disease, pulmonary disease, smoking, renal failure) (18), in-hospital laboratory results (completed blood count, markers previously shown to be associated with severity of illness at presentation such as D-dimer, C-reactive protein [CRP], troponin), imaging tests (using the first available of chest x-ray [CXR], CT chest, echocardiogram, and doppler ultrasound), medications, and clinical outcomes. Our proxies for disease severity included markers known to be associated with worse clinical outcomes: CRP, troponin, creatinine, D-dimer, abnormal CXR, and need for supplemental oxygen (19). Text mapping using natural language processing was used to characterize the findings on each imaging report.

### Patient Follow-up

Measurements began on the date of hospitalization for COVID-19 (the index date) and follow-up continued until death, discharge from hospital, or 30 days from the index date. This specification of the follow-up period was the same as that used in the in-hospital COVID-19 clinical trials (20-22). Our primary outcome was within hospital 30-day risk of death.

### Data analysis

Descriptive statistics were used to compare baseline characteristics between the two groups. The association between diabetes and death was measured using Poisson regression that include patient level characteristics including age, sex, admission location, comorbid conditions (heart failure, cerebrovascular disease [CVD], hypertension [HTN], COPD/asthma, smoking, and kidney disease, and proxies for illness severity (CRP, troponin, creatinine, D-dimer, and need for supplemental oxygen). A separate Poisson regression model was performed in the Ontario and Danish data and the results were meta-analyzed to provide an overall rate ratio for mortality

## Results

Our study included 1,438 hospitalized patients with COVID-19, including 1,133 (78.8%) from Ontario and 305 (21.2%) from Denmark. 480 patients (33.4%) had diabetes, 405 from Ontario and 75 from Denmark. In Ontario, people with diabetes were older and had more comorbidities (e.g., chronic kidney disease, cardiovascular disease, heart failure and hypertension) than those who did not have diabetes (Table 1a). They were also more likely to be from a long-term care home and have higher severity of illness at presentation to hospital (e.g., higher CRP and D-dimer, abnormal chest x-ray) (Table 2a). They also had higher troponin and creatinine values compared with patients without diabetes. In Denmark, patients with diabetes also were older and had more comorbidities than those without diabetes (Table 1b). They had a higher creatinine and troponin at time of presentation as well. In Denmark, CRP and D-dimer levels were similar between those with diabetes and those without, and both groups were equally likely to have an abnormal CXR (Table 2b).

**Table 1a.** Baseline characteristics of included patient population (Ontario)

| Variable | Strata | Diabetes (n=405) | No Diabetes (n=728) |
| --- | --- | --- | --- |
| **Age (Mean, SD)** | Age (Mean SD) | 69.9 (14.0) | 62.8 (19.8) |
| **Age group (n, %)** | <50 years | 32 (7.9) | 190 (26.0) |
|  | 50-59 years | 54 (13.3) | 114 (15.7) |
|  | 60-69 years | 98 (24.2) | 124 (17.0) |
|  | 70-79 years | 112 (27.7) | 124 (17.0) |
|  | 80-89 years | 82 (20.2) | 117 (16.1) |
|  | 90+ Years | 27 (6.7) | 59 (8.1) |
| **COVID Positivity Date** | On/Before April 25th, 2020 | 243 (60) | 421 (57.8) |
|  | After April 25<sup>th</sup>, 2020 | 162 (40) | 307 (42.2) |
| **Nosocomial COVID (%)** |  | 55 (13.6) | 86 (11.8) |
| **Sex (n, %)** | Female | 171 (42.2) | 308 (42.3) |
| **Limited English Proficiency (%)** |  | 50 (12.3) | 76 (10.4) |
| **Admission Location (n, %)** | Home | 252 (62.2) | 460 (63.2) |
|  | LTC | 91 (22.5) | 120 (16.5) |
|  | Hospital (Transfer) | 49 (12.1) | 79 (10.9) |
|  | No fixed home address | 13 (3.2) | 69 (9.5) |
| **Comorbidities (n, %)** | Hypertension | 316 (78.0) | 326 (44.8) |
|  | Heart Failure | 85 (20.9) | 76 (10.4) |
|  | Smoking | 97 (23.9) | 148 (20.3) |
|  | Cardiovascular Dx | 130 (32.1) | 135 (18.5) |
|  | COPD | 85 (20.9) | 122 (16.8) |
|  | CKD | 112 (27.7) | 59 (8.1) |
| **Creatinine Group, μmol/L (%)** | 0-100 | 153 (37.8) | 466 (64.0) |
|  | 101-200 | 138 (34.1) | 142 (19.5) |
|  | >200 | 78 (19.3) | 37 (5.1) |
|  | Missing | 36 (8.9) | 83 (11.4) |
| **A1C, % (Mean, SD)** |  | 7.844 (2.3) | 5.618 (0.4) |
|  | NA (%) | 243 (60.0) | 663 (91.1) |
| **Lymphocyte, $10^9$ cells/L (Mean, SD)** |  | 1.133 (0.8) | 1.196 (0.8) |
|  | NA (%) | 43 (10.6) | 80 (10.9) |
| **Ferritin, μg/L (Mean, SD)** |  | 937.932 (1443.5) | 972 (2096.2) |
|  | NA (%) | 198 (48.9) | 313 (42.9) |
| **Presence of CXR (%)** |  | 345 (85.2) | 597 (82.0) |

SD = standard deviation; LTC = long term care; COPD = chronic obstructive pulmonary disease; CKD = chronic kidney disease; WBC = white blood cell; CXR = chest x-ray

**Table 1b.** Baseline characteristics of included patient population (Denmark).

| **Variable** | **Strata** | **Diabetes (n=75)** | **No Diabetes (n=230)** |
| --- | --- | --- | --- |
| **Age (Mean, SD)** | Age (Mean SD) | 70.3 (13.0) | 68.1 (13.9) |
| **Age group (n, %)** | 50-59 | 16 (21.3) | 67 (29.1) |
|  | 60-69 | 16 (21.3) | 51 (22.2) |
|  | 70-79 | 23 (30.7) | 52 (22.6) |
|  | 80-89 | 17 (22.7) | 49 (21.3) |
|  | 90+ | 3 (4.0) | 11 (4.8) |
| **COVID Positivity Date** | On/Before April 25th, 2020 | 47 (62.7) | 139 (60.4) |
|  | After April 25<sup>th</sup>, 2020 | 28 (37.3) | 91 (39.6) |
| **Nosocomial COVID (%)** |  | 14 (18.7) | 36 (15.7) |
| **Sex (n, %)** | Female | 29 (38.7) | 102 (44.3) |
| **Admission Location (n, %)** | Home | 61 (81.3) | 194 (84.3) |
|  | LTC/Hospital | 14 (18.7) | 36 (15.7) |
| **Comorbidities (n, %)** | Hypertension | 47 (62.7) | 126 (54.8) |
|  | Heart Failure | 10 (13.3) | 24 (10.4) |
|  | Smoking | 43 (57.3) | 127 (55.2) |
|  | Cerebrovascular Dx | 14 (18.7) | 37 (16.1) |
|  | COPD | 13 (17.6) | 33 (14.3) |
|  | CKD | 17 (22.7) | 29 (12.6) |
| **Creatinine Group, μmol/L (%)** | 0-100 | 46 (61.3) | 179 (77.8) |
|  | 101-200 | 21 (28.0) | 32 (13.9) |
|  | >200 | 8 (10.7) | 16 (7.0) |
| **A1C, mmol/mol (Mean, SD)** |  | 55.1 (19.8) | 40.9 (6.1) |
| **Lymphocyte, $10^9$ cells/L (Mean, SD)** |  | 1.6 (1.7) | 1.2 (1.0) |
| **Ferritin, μg/L (Mean, SD)** |  | 936.7 (1216.7) | 947.3 (1064.2) |
| **Presence of CXR (%)** |  | 71 (94.7) | 218 (94.8) |

SD = standard deviation; LTC = long term care; COPD = chronic obstructive pulmonary disease; CKD = chronic kidney disease; WBC = white blood cell; CXR = chest x-ray

**Table 2a.** Markers of Illness Severity (Ontario)

| Variable | Strata | Diabetes (n=405) | No Diabetes (n=728) |
| --- | --- | --- | --- |
| **CRP Group, mg/L (%)** | 0-14.9 | 23 (5.7) | 75 (10.3) |
|  | 15-100 | 105 (25.9) | 167 (22.9) |
|  | >100 | 90 (22.2) | 138 (18.9) |
|  | Missing | 187 (46.2) | 348 (47.8) |
| **Median CRP (IQR)** |  | 79.3 (37.2 – 151.1) | 65.5 (20 – 135.3) |
| **D-Dimer Group** | <ULN | 26 (6.4) | 74 (10.2) |
|  | ULN-2*ULN | 61 (15.1) | 111 (15.2) |
|  | >2*ULN | 137 (33.8) | 199 (27.3) |
|  | Missing | 181 (44.7) | 344 (47.3) |
| **Troponin Group** | <ULN | 151 (37.3) | 294 (40.4) |
|  | ULN-2*ULN | 66 (16.3) | 89 (12.2) |
|  | >2*ULN | 84 (20.7) | 99 (13.6) |
|  | Missing | 104 (25.7) | 246 (33.8) |
| **Oxygen requirement (%)** |  | 57 (14.1) | 98 (13.5) |
|  | NA (%) | 303 (74.8) | 537 (73.7) |
| **Presence of ABG** |  | 72 (17.8) | 102 (14.0) |
| **CXR Results** | Normal CXR (%) | 59 (14.6) | 137 (18.8) |
|  | Abnormal CXR (%) | 227 (56.1) | 371 (50.9) |
|  | Missing CXR (%) | 61 (15.1) | 134 (18.4) |
|  | Unknown CXR (%) | 39 (9.6) | 63 (8.7) |

CRP = c-reactive protein; IQR = interquartile range; ULN = upper limit of normal; ABG = arterial blood gas; CXR = chest x-ray

**Table 2b.**
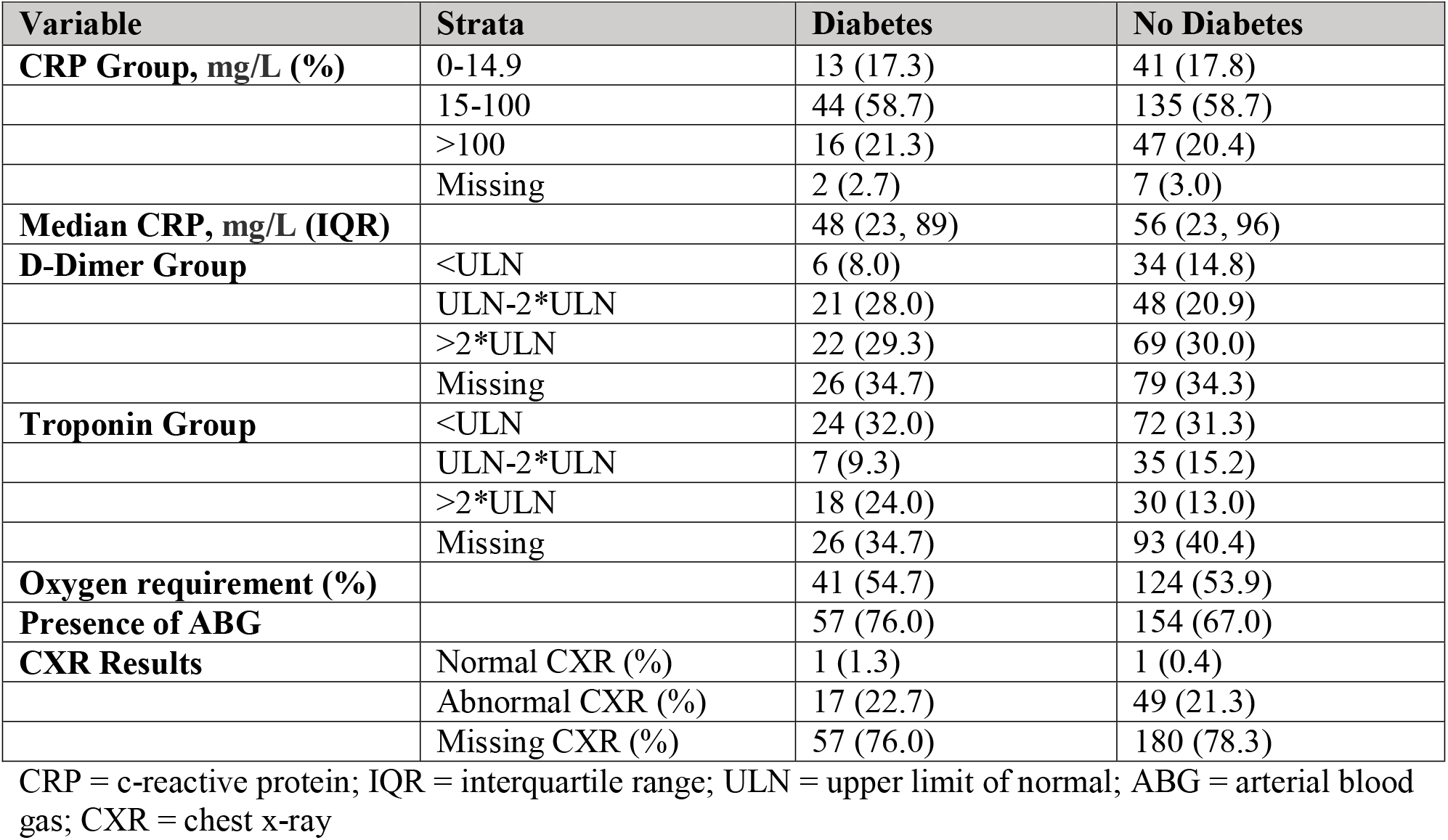
Markers of Illness Severity (Denmark)

| Variable | Strata | Diabetes | No Diabetes |
| --- | --- | --- | --- |
| **CRP Group, mg/L (%)** | 0-14.9 | 13 (17.3) | 41 (17.8) |
|  | 15-100 | 44 (58.7) | 135 (58.7) |
|  | >100 | 16 (21.3) | 47 (20.4) |
|  | Missing | 2 (2.7) | 7 (3.0) |
| **Median CRP, mg/L (IQR)** |  | 48 (23, 89) | 56 (23, 96) |
| **D-Dimer Group** | <ULN | 6 (8.0) | 34 (14.8) |
|  | ULN-2*ULN | 21 (28.0) | 48 (20.9) |
|  | >2*ULN | 22 (29.3) | 69 (30.0) |
|  | Missing | 26 (34.7) | 79 (34.3) |
| **Troponin Group** | <ULN | 24 (32.0) | 72 (31.3) |
|  | ULN-2*ULN | 7 (9.3) | 35 (15.2) |
|  | >2*ULN | 18 (24.0) | 30 (13.0) |
|  | Missing | 26 (34.7) | 93 (40.4) |
| **Oxygen requirement (%)** |  | 41 (54.7) | 124 (53.9) |
| **Presence of ABG** |  | 57 (76.0) | 154 (67.0) |
| **CXR Results** | Normal CXR (%) | 1 (1.3) | 1 (0.4) |
|  | Abnormal CXR (%) | 17 (22.7) | 49 (21.3) |
|  | Missing CXR (%) | 57 (76.0) | 180 (78.3) |

CRP = c-reactive protein; IQR = interquartile range; ULN = upper limit of normal; ABG = arterial blood gas; CXR = chest x-ray

In both Ontario and Denmark, people with diabetes were more likely to receive an antibiotic and less likely to receive an anti-viral medication (Tables 3a and 3b). In Ontario, steroid use was uncommon and occurred at a similar frequency for the two groups. There was also no clear difference in the number of patients with and without diabetes who received a CT thorax, echocardiogram, or doppler ultrasound of the lower extremities (Table 3a). In Denmark, all patients received an echocardiogram, none received a biologic, and data on doppler studies and steroid administration was not collected (Table 3b).

**Table 3a.** Patterns of care in hospital (Ontario).

| **Variable** | **Diabetes** | **No Diabetes** |
| --- | --- | --- |
| **CT Thorax (%)** | 97 (23.9) | 155 (21.3) |
| **Echocardiogram (%)** | 43 (10.6) | 59 (8.1) |
| **Doppler ultrasound (%)** | 47 (11.6) | 68 (9.3) |
| **Antibiotic received (%)** | 304 (75.1) | 492 (67.6) |
| **Antiviral received (%)** | 52 (12.8) | 114 (15.7) |
| **Steroid received (%)** | 92 (22.7) | 145 (19.9) |
| **Biologic received (%)** | 3 (0.7) | 9 (1.2) |

**Table 3b.** Patterns of care in hospital (Denmark)

| **Variable** | **Diabetes** | **No Diabetes** |
| --- | --- | --- |
| **CT Thorax (%)** | 14 (18.7) | 50 (21.8) |
| **Echocardiogram (%)** | 75 (100.0) | 230 (100.0) |
| **Antibiotic received (%)** | 63 (84.0) | 166 (72.2) |
| **Antiviral received (%)** | 12 (16.0) | 45 (19.6) |
| **Biologic received (%) - none** | 0 (0) | 0 (0) |

In Ontario, the crude mortality ratio for patients with diabetes, compared with non-diabetic, was 1.60 (95% CI 1.24 – 2.07). After adjustment for patient-level characteristics (Table 4), the mortality ratio was 1.19 [95% CI 0.86 – 1.66]. In the Danish patient population, the crude mortality ratio was 1.27 (95% CI 0.68 – 2.36), but after control of confounders the adjusted mortality ratio was 0.87 [95% CI 0.49 – 1.54] (Table 4). Meta-analyzing the two rate ratios from each region resulted in a crude mortality rate ratio of 1.55 (95% CI 1.22,1.96) and an adjusted mortality rate ratio of 1.11 (95% CI 0.84, 1.47).

**Table 4a.**
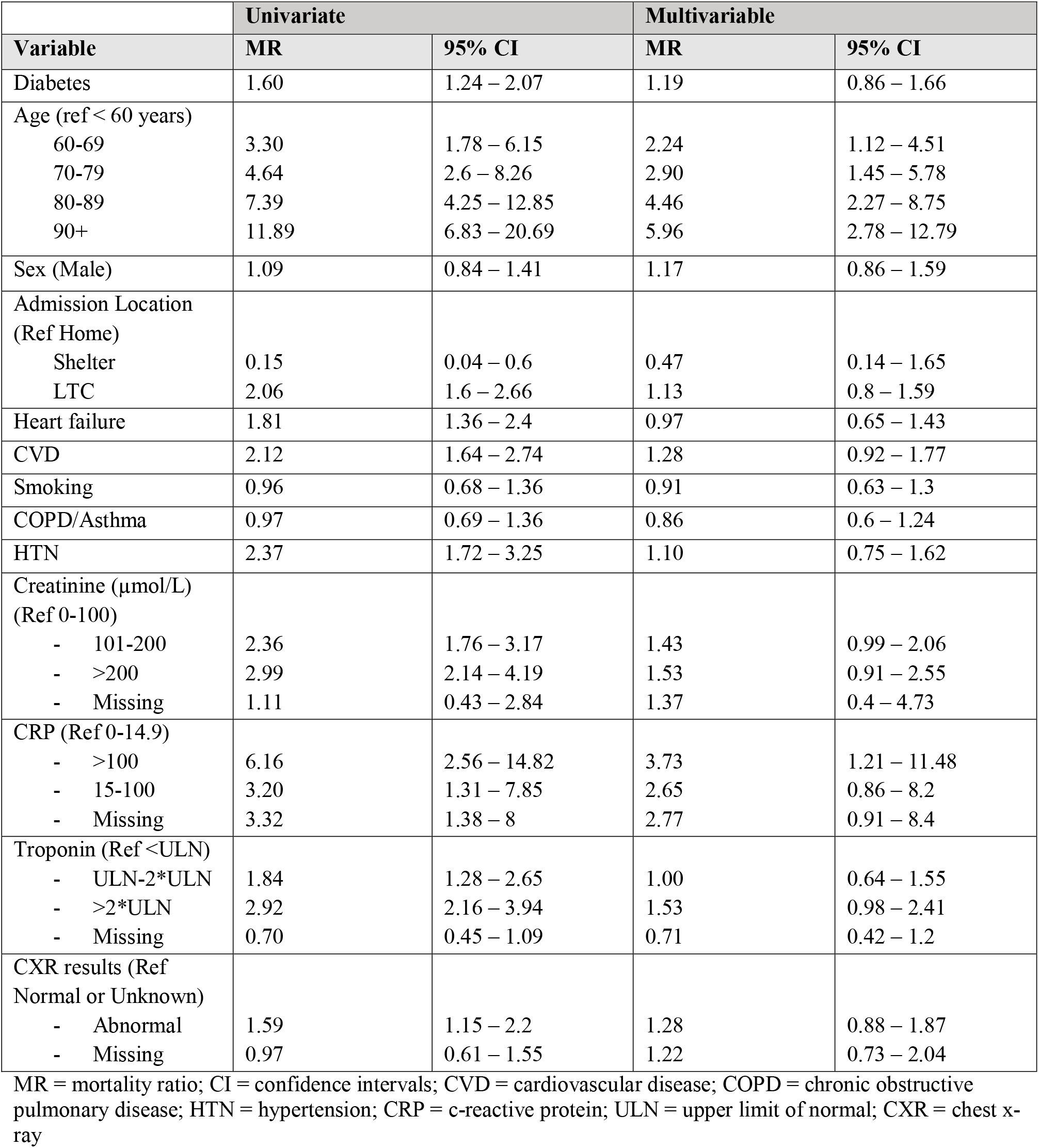
Univariate and multivariable model for the association between diabetes and death (Ontario).

| Variable | Univariate |  | Multivariable |  |
| --- | --- | --- | --- | --- |
|  | MR | 95% CI | MR | 95% CI |
| Diabetes | 1.60 | 1.24 – 2.07 | 1.19 | 0.86 – 1.66 |
| Age (ref < 60 years) |  |  |  |  |
| 60-69 | 3.30 | 1.78 – 6.15 | 2.24 | 1.12 – 4.51 |
| 70-79 | 4.64 | 2.6 – 8.26 | 2.90 | 1.45 – 5.78 |
| 80-89 | 7.39 | 4.25 – 12.85 | 4.46 | 2.27 – 8.75 |
| 90+ | 11.89 | 6.83 – 20.69 | 5.96 | 2.78 – 12.79 |
| Sex (Male) | 1.09 | 0.84 – 1.41 | 1.17 | 0.86 – 1.59 |
| Admission Location (Ref Home) |  |  |  |  |
| Shelter | 0.15 | 0.04 – 0.6 | 0.47 | 0.14 – 1.65 |
| LTC | 2.06 | 1.6 – 2.66 | 1.13 | 0.8 – 1.59 |
| Heart failure | 1.81 | 1.36 – 2.4 | 0.97 | 0.65 – 1.43 |
| CVD | 2.12 | 1.64 – 2.74 | 1.28 | 0.92 – 1.77 |
| Smoking | 0.96 | 0.68 – 1.36 | 0.91 | 0.63 – 1.3 |
| COPD/Asthma | 0.97 | 0.69 – 1.36 | 0.86 | 0.6 – 1.24 |
| HTN | 2.37 | 1.72 – 3.25 | 1.10 | 0.75 – 1.62 |
| Creatinine (µmol/L) (Ref 0-100) |  |  |  |  |
| - 101-200 | 2.36 | 1.76 – 3.17 | 1.43 | 0.99 – 2.06 |
| - >200 | 2.99 | 2.14 – 4.19 | 1.53 | 0.91 – 2.55 |
| - Missing | 1.11 | 0.43 – 2.84 | 1.37 | 0.4 – 4.73 |
| CRP (Ref 0-14.9) |  |  |  |  |
| - >100 | 6.16 | 2.56 – 14.82 | 3.73 | 1.21 – 11.48 |
| - 15-100 | 3.20 | 1.31 – 7.85 | 2.65 | 0.86 – 8.2 |
| - Missing | 3.32 | 1.38 – 8 | 2.77 | 0.91 – 8.4 |
| Troponin (Ref <ULN) |  |  |  |  |
| - ULN-2*ULN | 1.84 | 1.28 – 2.65 | 1.00 | 0.64 – 1.55 |
| - >2*ULN | 2.92 | 2.16 – 3.94 | 1.53 | 0.98 – 2.41 |
| - Missing | 0.70 | 0.45 – 1.09 | 0.71 | 0.42 – 1.2 |
| CXR results (Ref Normal or Unknown) |  |  |  |  |
| - Abnormal | 1.59 | 1.15 – 2.2 | 1.28 | 0.88 – 1.87 |
| - Missing | 0.97 | 0.61 – 1.55 | 1.22 | 0.73 – 2.04 |

MR = mortality ratio; CI = confidence intervals; CVD = cardiovascular disease; COPD = chronic obstructive pulmonary disease; HTN = hypertension; CRP = c-reactive protein; ULN = upper limit of normal; CXR = chest x-ray

**Table 4b.** Univariate and multivariable model for the association between diabetes and death (Denmark).

| Variable | Univariate |  | Multivariable |  |
| --- | --- | --- | --- | --- |
|  | MR | 95% CI | MR | 95% CI |
| Diabetes | 1.27 | 0.68 – 2.36 | 0.87 | 0.49 – 1.54 |
| Age (Ref <60) |  |  |  |  |
| 60-69 | 5.00 | 0.57 – 43.45 | 4.45 | 0.54 – 36.89 |
| 70-79 | 15.5 | 2.07 – 115.39 | 14.71 | 2.11 – 102.34 |
| 80-89 | 21.4 | 2.91 – 157.02 | 14.98 | 2.18 – 103.02 |
| 90+ | 29.6 | 3.72 – 235.95 | 17.75 | 2.28 – 138.06 |
| Sex (Male) | 2.05 | 1.07 – 3.95 | 1.34 | 0.68 – 2.63 |
| Admission Location (Ref Home) |  |  |  |  |
| LTC | 1.87 | 1 – 3.48 | 1.92 | 1.02 – 3.59 |
| Heart failure | 2.57 | 1.39 – 4.77 | 0.64 | 0.33 – 1.25 |
| CVD | 2.58 | 1.46 – 4.58 | 1.89 | 0.94 – 3.79 |
| Smoking | 1.12 | 0.63 – 2 | 0.68 | 0.32 – 1.47 |
| COPD/Asthma | 0.86 | 0.44 – 1.68 | 1.07 | 0.48 – 2.36 |
| HTN | 1.84 | 0.98 – 3.48 | 0.66 | 0.3 – 1.44 |
| Creatinine (µmol/L) (Ref 0-100) |  |  |  |  |
| - 101-200 | 4.25 | 2.27 – 7.93 | 3.96 | 1.8 – 8.68 |
| - >200 | 4.69 | 2.24 -9.81 | 2.17 | 0.92 – 5.09 |
| - Missing | 4.69 | 0.88 – 24.93 | 2.55 | 0.44 – 14.62 |
| CRP (Ref 0-14.9) |  |  |  |  |
| - >100 | 7.29 | 1.76 – 30.19 | 4.93 | 1.28 – 19 |
| - 15-100 | 3.17 | 0.76 – 13.11 | 2.61 | 0.71 – 9.59 |
| - Missing | 3.00 | 0.3 – 29.87 | 1.6 | 0.22 – 11.71 |
| Troponin (Ref <ULN) |  |  |  |  |
| - ULN-2*ULN | 3.81 | 0.95 – 15.25 | 1.52 | 0.39 – 5.95 |
| - >2*ULN | 6.00 | 1.7 – 21.19 | 1.00 | 0.28 – 3.62 |
| - Missing | 6.45 | 2 – 20.83 | 3.88 | 1.08 – 13.97 |
| CXR results (Ref Normal/Unknown) |  |  |  |  |
| - Abnormal | 0.48 | 0.11 – 2.07 | 3.15 | 0.97 – 10.2 |
| - Missing | 0.20 | 0.048 – 0.85 | 1.13 | 0.42 – 3.05 |

MR = mortality ratio; CI = confidence intervals; CVD = cardiovascular disease; COPD = chronic obstructive pulmonary disease; HTN = hypertension; CRP = c-reactive protein; ULN = upper limit of normal; CXR = chest x-ray

## Discussion

After control of confounding, diabetes was only weakly associated with within hospital 30-day risk of death, with no consistent pattern in Canada and Denmark. Multiple studies assessing whether diabetes was associated with an increased risk of mortality lacked information on severity of illness on presentation (e.g., chest x-ray findings) or lab values associated with a worse prognosis (e.g., troponin, d-dimer, creatinine). The strongest predictor of respiratory failure and mortality among COVID patients, aside from age, is illness severity at presentation (19). Studies that fail to account for these important confounders may overestimate the independent effect of diabetes on COVID outcomes.

Patients with diabetes tended to have both more severe illness (e.g., higher CRP, worse chest x-ray findings) and worse prognostic signs in both countries that we studied. It is unknown if patients with diabetes are sicker on presentation because of delays in seeking care causing selection bias, association between diabetes and other factors that may influence severity such as race or socioeconomic deprivation, or the direct influence of diabetes. The higher illness severity may explain why patients with diabetes were more likely to receive antibiotics while in hospital, though subsequent trials have identified that antibiotics are ineffective against COVID-19 (20). The higher use of antibiotics in patients with diabetes may also be explained by their inherent immune-compromised state and their susceptibility to bacterial co-infection.

Our study has important limitations. First, not all patients had a hemoglobin A1C measured and thus only a subset of patients have these data available. As a result, we were not able to identify severity of diabetes for some of the included patients. Second, we did not capture the proportion of patients with type 1 diabetes versus type 2 diabetes; we estimate that approximately 90% of patients had type 2 diabetes based on other published literature (25).

Third, we lacked data on other important comorbid conditions such as elevated BMI. A prior meta-analysis identified that the relative risk of mortality was 3.52 in those with BMI ≥ 25 and thus an important risk factor for COVID-19 outcomes (26). Residual confounding related to BMI would mean that we overestimated the relationship between diabetes and mortality, which was already weak after adjustment for confounders. Fifth, this study was conducted in two high-income countries (Denmark and Canada) with socialized healthcare systems; the results may thus not be generalizable to areas of low-income and/or private-paying systems. Unfortunately, our dataset lacked important social determinants of health such as income, race or ethnicity and education level. Sixth, our study was relatively small. Nonetheless, our finding that diabetes itself may not be a strong risk factor for death within 30 days of hospitalization warrants corroboration from other patient populations.

## Data Availability

All data produced in the present work are contained in the manuscript.

